# Becoming a resilient scientist series: An intervention program

**DOI:** 10.1101/2023.05.02.23289388

**Authors:** H. Anna Han, Ulrike Klenke, Laurie Chaikind McNulty, Annie Scheiner, Sharon L. Milgram

## Abstract

Compared to the general population, science trainees experience challenges and heightened stressors that often lead to adverse mental health outcomes. With COVID-19, the stressors of social distancing, isolation, truncated lab time, and uncertainty about the future have all likely exacerbated these issues. Now, more than ever, practical and effective interventions are vitally needed to address the core causes of stress among science trainees and increase their resilience. This paper introduces a new resilience program targeted to biomedical trainees and scientists – Becoming a Resilient Scientist Series (BRS), a multi-part workshop complemented by facilitated group discussions all aimed at bolstering resilience, particularly in the context of academic and research environments. To assess the program’s efficacy, participants completed resilience measures and related assessments before and after completing the series. The results demonstrate that BRS significantly enhances trainee resilience (primary outcome) and reduces perceived stress, anxiety, and work-related presenteeism, as well as increased adaptability, self-awareness, and self-efficacy (secondary outcomes). Furthermore, program participants reported a high level of satisfaction, a strong willingness to recommend the program to others, and perceived positive changes in their resilience skills. To the best of our knowledge, this is the first resilience program designed explicitly for biomedical trainees and scientists, tailored to their unique professional culture and work environment.

## Introduction

### The mental health of science trainees

Science is hard. And this is not just a cliché or cultural myth – the data support it. In fact, science is so hard that 60% of students starting in science, technology, engineering, and mathematic disciplines (STEM) and pre-med major will change to non-STEM fields at a rate 2 times higher than other non-science fields [1–2]. Grades, by the same or comparable individuals, are substantially lower in science versus non-science courses [3].

But science is also hard for reasons other than the topic of study. After surviving the undergraduate math and science attrition effect, many of aspiring scientists face mental health crises in the field caused by a myriad of academic stressors. These include imposter fears (vs. imposter syndrome, as the term syndrome seems to denote something is wrong with the individuals or that it is abnormal), isolation, constant looming deadlines, navigating complex relationships with advisers, intense competition, lack of work-life balance, uncertain job prospects, and bullying and harassment. For example, a survey of 4,300 academic scientists worldwide reported that at least half struggled with self-reported depression and anxiety, and 67% reported witnessing bullying or harassment, with 43% directly experiencing either bullying or harassment [4]. This effect is most pronounced among graduate students and trainees who are at 6 times greater risk for depression and anxiety than the general population [5]. In *Nature*’s survey of doctoral students and their training experiences [6], 36% respondents reported seeking help for depression and anxiety that stem from their training. Almost half (45%) said their satisfaction with their PhD trajectory decreased as they progressed in their training.

With COVID-19, these negative effects were exacerbated by increased social isolation, truncated lab time, and financial stressors. In fact, in a survey of medical scientists including medical and graduate students, 23% of respondents considered leaving academia post COVID-19, due to lack of work-life balance [7]. This effect is even more pronounced amongst biomedical scientists [8]. A survey conducted in the Netherlands during the height of the pandemic from March to May 2020 revealed that 47% of PhD trainees were at risk for psychiatric disorder, and approximately 40% experienced severe burn-out symptoms [9]. As Chan et al. described it “…imagine the mental resilience needed to maintain focus on solving that equally important mystery in oncology, cardiology, neuroscience, or any other field that has been put on temporary hold due to the pandemic” [8].

Science, especially the biomedical sciences, has already been suffering a high attrition rate in PhD programs, and the attrition is even more pronounced with women and underrepresented minority (URM) trainees who have been leaving science and academia at a disproportionate rate [10–11]. Without practical and effective interventions to address the stresses experienced by biomedical trainees, which exacerbate mental health crises, the field will suffer a brain drain and lose talented future scientists and potential for innovation. Now, more than ever, practical and effective interventions are vitally needed to address the core causes of biomedical trainee stress and to increase resilience amongst trainee populations.

While many of the findings, both for scientists in general and biomedical trainees in particular, raise alarms and call for immediate intervention to help those in the scientific workforce pipeline, academia’s responses have been largely muted. *Inside Higher Ed* declared a mental crisis in graduate education and stated, “It is only with strong and validated interventions that academia will be able to provide help for those who are traveling through the bioscience workforce pipeline”[12]. However, available interventions are lacking, and most are not validated, especially for biomedical science trainees. In fact, the results from the Graduate Student Depression and Anxiety Survey led by Evans and colleagues to recommend National Institutes of Health’s (NIH) own train-the-trainer model, where faculty, administrative, and support staff are trained by mental health professionals to recognize and respond to the trainee’s needs, be adapted “to help today’s PhDs compete in the ‘vast and ever-changing job market’” [5].

Here, we propose that to alleviate the current mental health crisis within graduate education, an effective intervention needs to address the unique challenges of academia, target the sources of depression and anxiety, and increase trainees’ ability to cope with stressors and adversity. One promising approach is to provide tailored training that increases trainees’ resilience. For example, a systematic review of resilience training and interventions [13] revealed a large body of evidence highlighting the benefits of resilience training for mental health and well-being by mitigating the impact of stress and adversity. Additionally, in a recent study of women who are thriving in undergraduate STEM majors, resilience was identified as a common and integral trait that allowed them to succeed [14]. In exploring why trainees withdraw early from their biomedical PhD programs, Maher and colleagues [15] found that self-efficacy components related to resiliency played a role (see 16 for a broader analysis of reasons why women exit STEM majors).

### Resilience as an intervention?

Resilience is a form of mental and psychological strength that enables a person to adapt and adjust to difficult or stressful situations. According to the American Psychological Association, resilience is “the process of adapting well in the face of adversity, trauma, tragedy, threats, or significant sources of stress — such as family and relationship problems, serious health problems or workplace and financial stressors. It means ‘bouncing back’ from difficult experiences” [17]. Because of their ability to adapt, resilient individuals tend to better regulate their behaviors, have a more optimistic outlook with greater life satisfaction, maintain positive self-views. They are also less likely to be depressed and anxious [18–20]. Resilient individuals are also less likely to engage in presenteeism – that is, they are more likely to remain fully functioning in the workplace rather than working while distracted [21] – and they also avoid other self-defeating work behaviors [22–23]. Importantly, resilience is malleable and can be learned and nurtured [24].

Although resilience training has seldom been attempted in academic settings, resilience training intervention has been shown to be effective in workplace and military settings (e.g., U.S. Army’s Ready and Resilient Campaign). For example, resilience training increased positive affect and a sense of well-being while it decreased depression and anxiety [25–26]. Moreover, such training has been linked to increased work performance [26] and organizational commitment [27]. An in-depth systematic review of resilience training interventions in the workplace demonstrated that such trainings generally improved personal resilience, mental health, and subjective well-being of participants, and had other tangible benefits including improved performance and improved psychosocial outcomes such as increased self-efficacy and optimism [26]. It has been estimated that the cost-savings for resilience intervention training is $1,846 per person over 8-weeks due to the reduction in stress tied to depression or trait anxiety, as well as increased presenteeism of workers [28].

Given the constant stressors and rejection faced in scientific academic settings, and the high levels of depression and anxiety experienced by graduate trainees, there is a clear need for resilience training aimed at increasing an individual’s ability to adapt in the face of adversity and “bounce back,” as well as decrease stress, depression, and anxiety, and other self-defeating work behaviors tailored especially to biomedical trainees. The objective of the current paper is to investigate whether a novel resilience program tailored for biomedical trainees and scientists can enhance their resilience, and hence, their persistence in science.

### Becoming a resilient scientist (BRS): The intervention program

The Becoming a Resilient Scientist Series (BRS) originally evolved from several standalone webinars conducted by the NIH Office of Intramural Training and Education (OITE) to help trainees manage stress. As the demand for these webinars increased at the start of the pandemic, the questions and responses from the trainees clearly indicated that a more cohesive and comprehensive set of lectures and intervention was needed. Thus, the BRS was created in 2020 as a step toward meeting the needs of the trainees and helping alleviate and address common stressors and increase resilience for those pursuing science.

The BRS program employs multimodal cognitive-behavioral concepts that emphasizes community, mindfulness, self-compassion, and cognitive behavioral changes – all of which are thought to increase resilience. The intervention focuses on several broad themes across all sessions, including the importance of learning and practicing resilience skills and the ongoing nature of resilience-building, cultural awareness, the role of identity in the scientific community, and the critical role of community support. The program also emphasizes the potential benefits of therapy and mental health care while acknowledging possible barriers, such as stigma, cost, and the fear of losing productive work time. The program’s goal is to help participants identify and replace maladaptive coping strategies with more adaptive behaviors that support self-efficacy and persistence in STEM fields.

The program is a series of five two-hour workshops; each workshop is followed by an optional one hour facilitated small-group discussion. The five units are separated by one-to two-week intervals, to enable trainees to learn the content, process it on their own, and explore it with their peers in the facilitated small-group discussion. Each workshop and related discussion session can be a stand-alone, but trainees who attend the entire program can refine and integrate the insights and skills they have learned as material is reintroduced and reiterated throughout the series.

The BRS comprises five parts, each designed to benefit trainees in research and academic settings (see S1 for a full description of each session). Part one serves as the program’s foundation, addressing well-being practices, emotional literacy, and the development of a growth mindset. Trainees learn how to effectively cope with setbacks and disappointments by fostering resilience through habits like self-care, mindfulness, journaling, and seeking community support. Part two centers on countering cognitive distortions and imposter fears, offering strategies to combat negative self-talk and cultivate a growth mindset. Part three emphasizes self-advocacy and effective communication in academic and research hierarchies, teaching trainees how to set boundaries, communicate expectations, and address difficult issues. It particularly acknowledges the importance of mentorship, especially for marginalized trainees. Part four addresses the challenges of receiving feedback and staying receptive to it, and part five delves into effective mentoring and relationship management, with a focus on improving interactions with principal investigators, seeking additional mentors, and addressing toxic environments.

In the current paper, we evaluate the effectiveness of BRS as a resilience intervention program for biomedical science trainees who participated in the program during the COVID-19 pandemic. Given the severity of the pandemic, including widespread isolation, salient racial injustice issues, and concerns for various other mental health issues of the trainees at that time, a conscious and ethical decision was made to make the program open for all trainees who wanted to participate (vs. a waitlist). Although each workshop in the series could function as a stand-alone, we hypothesized that those who consistently participated in the program by attending more than half of the sessions would have more chances to integrate and practice the skills they learned, and therefore, would show greater increases in the primary outcome of resilience and associated secondary outcomes compared to those attending fewer than half of the sessions. Thus, the evaluation considers a “dose” effect independent of time on the primary and secondary outcomes, rather than a comparison to a waitlist control group who did not receive the training.

To that end, there are four major goals. First, we evaluate whether individuals who completed more than half (more than three sessions) of the BRS exhibited significantly higher increases in resilience levels compared to those who completed less than half. Second, since the program focused on themes of adjusting and adapting to adversity, stress, increasing self-awareness, believing in one’s ability to achieve goals, and other coping strategies, we expected corresponding changes on various correlates of resilience – decrease in perceived stress, depression, anxiety, work presenteeism, and increases in the ability to shift and persist during stressful events, self-efficacy, and self-awareness. Third, since the change in resilience should drive the changes in the secondary outcomes, we hypothesized that resilience should mediate the changes in the secondary outcome measures. Lastly, we describe participants’ reported satisfaction with program and whether self-reported changes differed for those completing the majority of sessions compared to those who completed fewer than half of the BRS program.

## Materials and methods

### Participants

Biomedical trainee participants were recruited from the NIH Intramural Research Program (IRP) and from various extramural institutions who were invited to participate in the program by the NIH’s OITE. For IRP trainees, the announcement for the program was made via OITE trainee listservs, and all IRP trainees (from postbaccalaureate to postdocs) were invited to participate. The trainees from extramural institutions were recruited by their institution, various listservs, and social media, and similar to NIH trainees, ranged from undergraduate to postdocs and medical students.

### Procedure

The current evaluation of the BRS program was implemented in two rounds, with the first round (BRS1) held from January to May 2021 across six sessions and the second round (BRS2) was held from September 2021 to December 2021 across five sessions. The workshop component was held once every 3 weeks for the first round (BRS1) and every 2 weeks for the second round (BRS2) via Zoom, and the small group discussions were held a week later. Trainees participated in the optional small group discussion sessions at their institution via Zoom or at one of the open sessions hosted at NIH via Zoom. Small discussion sessions were led by trained facilitators with a discussion guide. Because the content of the additional session in BRS1 was folded into other sessions of BRS2 and because there were no differences in data or attendance rates between the two sessions, the two rounds of BRS were collapsed into one.

Each webinar session had, on average, 363 attendees, with an additional 350 watching the recording. In addition, on average, 371 trainees attended the small group discussion sessions.

Prior to the start of the first BRS session, all participants who logged into the webinar were asked to complete a pre-program survey assessing current resilience levels (primary outcome) and secondary outcomes, such as perceived stress, anxiety and depression levels, work presenteeism, their current ability to shift and persist during stressful events, self-awareness, and self-efficacy (see measures below). Following completion of the BRS program, all participants who attended at least one workshop received an email with a link to the post-program survey. The post-program survey was identical to the pre-program survey but also included questions regarding program satisfaction, self-perceived changes, how many workshops were attended, demographics, and open-ended comments. The NIH Institutional Review Board granted an IRB exemption for this study, and the participants were provided with an online written consent at the beginning of each survey. The average length of time between pre-and post-survey was four months (the duration of BRS series), and the post-survey was open for 6 weeks after the conclusion of the series.

## Measures

### Program participation

At the post-program assessment, participants reported on the workshops they attended. Those who participated in more than three workshops were classified as “consistent attenders,” having attended more than half of the BRS program sessions; otherwise, participants were classified as “inconsistent attenders.”

### Primary outcome: Resilience

Resilience was measured by the 10-item Connor-Davidson Resilience Scale (CD-RISC-10) [29]. Participants were asked to rate the frequency (1 =’not true at all’ to 5 =’true nearly all the time’) in which they endorsed resilience related thoughts, beliefs, or behaviors in the last month (e.g., “I am able to adapt when changes occur”). In the current study, the items had good internal consistency with Cronbach’s α =0.83 (pre-program assessment) and 0.87 (post-program assessment). The ten items were averaged to create a score from 1-5, where higher scores indicate greater resilience.

### Secondary Outcomes

In the interest of survey response time, most secondary outcomes were assessed using abbreviated versions of well-validated measures of distress, well-being, and work engagement.

Perceived stress was measured by a subset of 4 items from the 10-item Perceived Stress Scale (PSS) [30] that focused on the control aspects of stress. Participants were asked to rate the frequency in which they experience stress-related feelings and thoughts (“how often have you felt that you were unable to control the important things in your life”) in the past month. Participants used a scale ranging from “never” (= 1) to “very often” (= 5). In the current study, the four items had acceptable internal consistency with Cronbach’s α =0.73 (pre-program assessment) and 0.77 (post-program assessment). The four items were averaged to create a score from 1-5, where the higher number indicates greater perceived stress.

Anxiety was measured by a subset of 3 items from the 7-item Generalized Anxiety Disorder Scale (GAD-7) [31], that focused on non-physical symptom items. Participants were asked to rate the frequency in which they experienced various anxiety symptoms (“Not being able to stop or control worrying”) in the past two weeks on a scale ranging from “not at all” (= 1) to “almost every day” (= 4). In the current study, the items had good internal consistency with Cronbach’s α = 0.76 (pre-program assessment) and 0.77 (post-program assessment). The three items were averaged to create a score from 1-4, where the higher number indicates greater anxiety levels.

Depression was measured by a subset of 4 items of the 9-item Patient Health Questionnaire (PHQ-9) [32] that focused on general non-clinical depressive symptoms. Participants were asked to rate the frequency in which they experienced various depressive symptoms (“feeling down, depressed, or hopeless”) in the past two weeks on a scale ranging from “not at all” (= 1) to “almost every day” (= 4). In the current study, the items had good internal consistency with Cronbach’s α =0.81 (pre-program assessment) and 0.79 (post-program assessment). The four items were averaged to create a score from 1-4, where the higher number indicates greater (non-clinical) depression levels.

Presenteeism was measured by the 6-item Job Stress Related Presenteeism Scale (JSRP) [33]. Participants were asked to rate the extent to which they had engaged in thoughts or behaviors related to presenteeism (“I’m unable to concentrate on my job because of work-related stress”) on a scale ranging from “never” (= 1) to “all the time” (= 5). In the current study, the items had good internal consistency with Cronbach’s α =0.83 (pre-program assessment) and 0.85 (post-program assessment). The six items were averaged to create a score from 1-5, where the higher number indicates greater job stress-related presenteeism.

The ability to shift and persist during stressful times was measured by the 14-item Shift-and-Persist Scale [34]. Participants were asked to rate the extent to which various statements describe them (“When something stressful happens in my life, I think about what I can learn from the situation”) on a scale of “not at all” (= 1) to “a lot” (= 4). In the current study, the items had good internal consistency with Cronbach’s α =0.82 (pre-program assessment) and 0.82 (post-program assessment). Excluding 6 distractor items, four items were average to create a shift sub-scale score from 1-4, and four items were average for a persist sub-scale score from 1-4. The higher scores indicate a greater ability to shift and/or persist during stressful times.

The belief in one’s ability to achieve their goals in the face of adversity was measured by the 8-item New General Self Efficacy Scale [35]. Participants were asked to indicate their agreement with statements such as, “I will be able to successfully overcome many challenges,” on a scale from “strongly disagree” (= 1) to “strongly agree” (= 5). In the current study, the items had good internal consistency with Cronbach’s α =0.89 (pre-program assessment) and 0.89 (post-program assessment). The eight items were averaged to create a score from 1-5. The higher scores indicate greater self-efficacy or the belief that one can overcome obstacles and achieve goals.

The awareness and reflection of one’s internal states with attention to learning and self-awareness at work were measured by a subset of items from the Self-Awareness Outcomes Questionnaire (SAOQ) [36], specifically the reflective self-development (RSD) and proactive at work (PRO) subscales. Because BRS focused on self-development and being proactive at work, the other subscales, acceptance and emotional costs, were not included as it may not be relevant to increased resilience. Participants were asked to rate the frequency in which they endorse statements such as “I focus on ways of amending my behavior that would be useful” on a scale of “never” (= 1) to “almost always” (= 5). In the current study, the items had good internal consistency with Cronbach’s α = 0.85 (pre-program assessment) and 0.87 (post-program assessment). Each subscale item was averaged to create a score from 1-5. The higher scores indicate greater self-awareness and reflection of oneself and self-awareness at work.

### Post Program Questions: Program Satisfaction, Perceived Changes, and Demographics

During the post-program survey, participants were asked to evaluate the BRS program with respect to overall satisfaction, likelihood they would recommend the training to a friend or colleague, and whether they found the program valuable on 5-point Likert scales.

To assess self-perceived changes, participants were asked during the post-program survey if they had become more resilient, better scientists, managed conflict and stress better, and if they had gained important skills on a 5-point Likert scale, ranging from 1-5 (e.g., “Since participating in the resilience series, I have become more resilient in my work and/or life.”). Furthermore, participants assessed their perceived knowledge on how to be a resilient scientist before and after participating in the program.

To assess program’s impact on different demographic groups, participants self-reported their gender identity and race/ethnicity. The gender identity options were man, woman, nonbinary, transgender, other, and prefer not to say. The race/ethnicity options were White, Asian, Black/African-American, Hispanic/Latino/Spanish, American Indian/Alaska Native, Native Hawaiian/Pacific Island, other, and prefer not to say. For both measures, participants could select more than one option, and those who selected multiple race/ethnic identities were coded as multi-racial and those who selected categories other than man and woman were coded as other.

## Data analysis

All analyses were conducted using SPSS. Prior to addressing study aims, we assessed whether there observed differences across trainee populations (i.e., NIH intramural trainees and extramural trainees) and BRS round (i.e., BRS1 and BRS2). There were no differences observed with regards to consistent participation, primary and secondary outcomes, and program satisfaction/perceived changes. Thus, data were collapsed across trainee populations and BRS rounds.

To address aims 1 and 2, we matched pre-and-post surveys and conducted a paired sample t-test to assess pre-program changes in resilience (primary outcome) and secondary outcomes at the post-program assessment. Furthermore, an independent sample t-test was used to explore the impact of consistent and inconsistent attendance on primary and secondary outcomes at the post-program assessment.

To address aim 3, we conducted a bootstrapped mediational analysis [37] to test indirect effect of consistent and inconsistent attendance on the secondary outcomes mediated by resilience.

To address aim 4, we constructed descriptive statistics for program satisfaction assessments and reported the percent rating the program ‘good’ (= 3) to ‘excellent’ (= 5). Independent t-tests were conducted to assess differences in program satisfaction and self-perceived changes following program completion between those who attended the program workshops consistently and those who were inconsistent attenders.

Given the attrition of females and underrepresented minorities in science, we further explored how trainee’s gender and race may influence the effects of BRS as an ancillary analysis.

## Results

### Participant characteristics

A total of 625 trainees completed the post-program survey across the two rounds of BRS. The analysis excluded three participants who identified as a facilitator or faculty, as the goal of this study was to look at the impact of the program on trainees. There were 440 females (70%), 154 males (25%), and 31 other/unknown (5%) gender. Two hundred and ninety-five trainees identified as white (47%), 308 minority/multiracial (49%), and 11 did not specific their race or ethnicity. Of those, using a unique self-guided ID, we were able to match up a total of 341 participants with their pre-and post-program surveys (216 in Spring 2021; 125 in Fall 2021). In the pre-and-post matched sample, there were 255 females (74.8%), 75 males (22%), and 11 other/unknown (3.3%). One hundred ninety-five trainees identified as white (57.2%), 142 as minority/multiracial (41.6%), and 4 (1.2%) did not specify their race or ethnicity.

### Aim 1: Change in resilience pre-to post-BRS program

To address aim 1, we conducted a paired sample t-test to assess pre-program changes in resilience at the post-program assessment. As predicted, there was a significant increase in RISC-10 resilience scores pre vs. post BRS program participation, (*M*_pre_ = 2.47 vs. *M*_post_ =2.80), such that participants resilience increased post BRS participation (Table 1).

**Table 1:** Pre-and-post program matched sample changes.

| Measure | Pre BRS Mean (SD) | Post BRS Mean (SD) | T-test, Cohen's d |
| --- | --- | --- | --- |
| **Resilience (RISC-10)** | 2.47 (0.57) | 2.80 (0.53) | $t(340) = 13.42$, $p < 0.001$, $d = 0.73$ |
| **Perceived Stress (Modified PSS)** | 1.99 (0.67) | 1.53 (0.68) | $t(340) = 11.94$, $p < 0.001$, $d = 0.65$ |
| **Anxiety (Modified GAD)** | 2.60 (0.75) | 2.28 (0.72) | $t(340) = 9.56$, $p < 0.001$, $d = 0.52$ |
| **Depressions**<br>**(Modified PHQ)** | 2.23 (0.79) | 1.97 (0.71) | $t(339) = 7.29, p < 0.001, d = 0.40$ |
| **Work Presenteeism**<br>**(JSRP)** | 2.53 (0.69) | 2.31 (0.67) | $t(339) = 6.39, p < 0.001, d = 0.35$ |
| **Shift &**<br>**Persist** | 2.92 (0.68)<br>3.11 (0.59) | 3.22 (0.62)<br>3.37 (0.58) | $t(339) = 9.90, p < 0.001, d = 0.54$<br>$t(339) = 8.39, p < 0.001, d = 0.45$ |
| **Self-Efficacy** | 3.85 (0.60) | 4.07 (0.53) | $t(338) = 8.03, p < 0.001, d = 0.44$ |
| **Self-Awareness:**<br>**Proactive at Work**<br>**&**<br>**Reflective Self-**<br>**Development** | 3.64 (0.53)<br>3.79 (0.54) | 3.83 (0.53)<br>3.99 (0.50) | $t(338) = 6.46, p < 0.001, d = 0.35$<br>$t(338) = 8.34, p < 0.001, d = 0.45$ |

Unsurprisingly, the matched pre-and-post sample had less than 14% individuals who attended 3 or less sessions and coded as “inconsistent attenders.” To explore the effect of consistent vs. inconsistent attendance on resilience more robustly, we used the post-program only sample and conducted an independent sample t-test to assess whether the resilience score differed for those who attended the BRS program consistently as compared to those who inconsistent attenders. As expected, we observed significantly higher RISC-10 resilience scores for consistent attenders (*M* = 2.82, SD = 0.50) vs. inconsistent attenders (*M* = 2.69, *SD* = 0.52), *t*(623) = 2.51, *p* = .012, *d* = 0.26) (see Table 2).

**Table 2:** Post-program scores by consistent vs. inconsistent attenders.

| **Measure** | **Consistent**<br>**Attenders**<br>**Mean (SD)** | **Inconsistent**<br>**Attenders**<br>**Mean (SD)** | **T-test, Cohen's d** |
| --- | --- | --- | --- |
| **Resilience (RISC-10)** | 2.82(0.50) | 2.69 (0.52) | $t(623) = 2.51, p = 0.012, d = 0.26$ |
| **Perceived Stress**<br>**(Modified PSS)** | 1.53(0.67) | 1.76 (0.68) | $t(622) = 3.14, p = 0.002, d = 0.33$ |
| **Anxiety (Modified**<br>**GAD)** | 2.25(0.71) | 2.30 (0.76) | $t(622) < 1, p = \text{NS}$ |
| **Depression**<br>**(Modified PHQ)** | 1.96 (0.79) | 2.08 (0.79) | $t(622) = 1.66, p = 0.098, d = 0.17$ |
| **Work Presenteeism**<br>**(JSRP)** | 2.28 (0.65) | 2.37 (0.66) | $t(622) < 1.5, p = \text{NS}$ |
| **Shift & Persist** | 3.23 (0.59)<br>3.40 (0.57) | 3.20 (0.62)<br>3.27 (0.73) | $t(622) < 1, p = \text{NS}$<br>$t(622) = 2.06, p = 0.039, d = 0.21$ |
| **Self-Efficacy** | 4.11 (0.52) | 3.95 (0.60) | $t(621) = 2.72, p = 0.007, d = 0.28$ |
| **Self-Awareness: Proactive at Work & Reflective Self-Development** | 4.04 (0.50)<br>3.85 (0.53) | 3.89 (0.59)<br>3.75 (0.59) | $t(622) = 1.81, p = 0.070, d = 0.19$<br>$t(622) = 2.70, p = 0.007, d = 0.28$ |

### Aim 2: Change in stress, self-efficacy, self-awareness, and persistence

With respect to secondary outcomes in the pre-and-post matched sample, we observed a significant decrease in perceived stress, anxiety, and depression and an increase in participants’ self-perceived ability to shift and persist, self-efficacy, self-awareness related to reflective self-development, and being proactive at work (Table 1). This suggests that those who consistently attend BRS from the start of the series are seeing significant changes in all secondary outcome measures.

Similarly, in the post-program sample, the consistent attenders were significantly lower in perceived stress, and higher ability to persist, self-efficacy, self-awareness related to reflective self-development. We saw marginal decreases in depression, self-awareness related to being proactive at work (Table 2). However, we did not see differences in anxiety, work presenteeism, or the ability to shift during stressful events.

### Aim 3: Mediation of resilience on consistent/inconsistent attendance and secondary outcomes

Mediational analyses based on 5,000 bootstrapped samples using bias-corrected and accelerated 95% confidence intervals [37–38] was conducted to test resilience (RISC-10) as a mediator of the relationship between attendance and secondary outcomes (Fig 1). In these analyses, mediation is significant when confidence intervals for the indirect effect do not include 0. As expected, resilience fully mediated depression, the ability to persist, self-efficacy, and perceptions of being proactive at work as well as reflective self-development components of self-awareness (Tables 3 and 4). It also partially mediated perceived stress. These results indicate that the increase in resilience by consistent attenders also account for the various secondary outcomes, including reduction in perceived stress, depression, and the increase in the ability to persist during stressful times, self-efficacy, and self-awareness.

**Fig 1.**
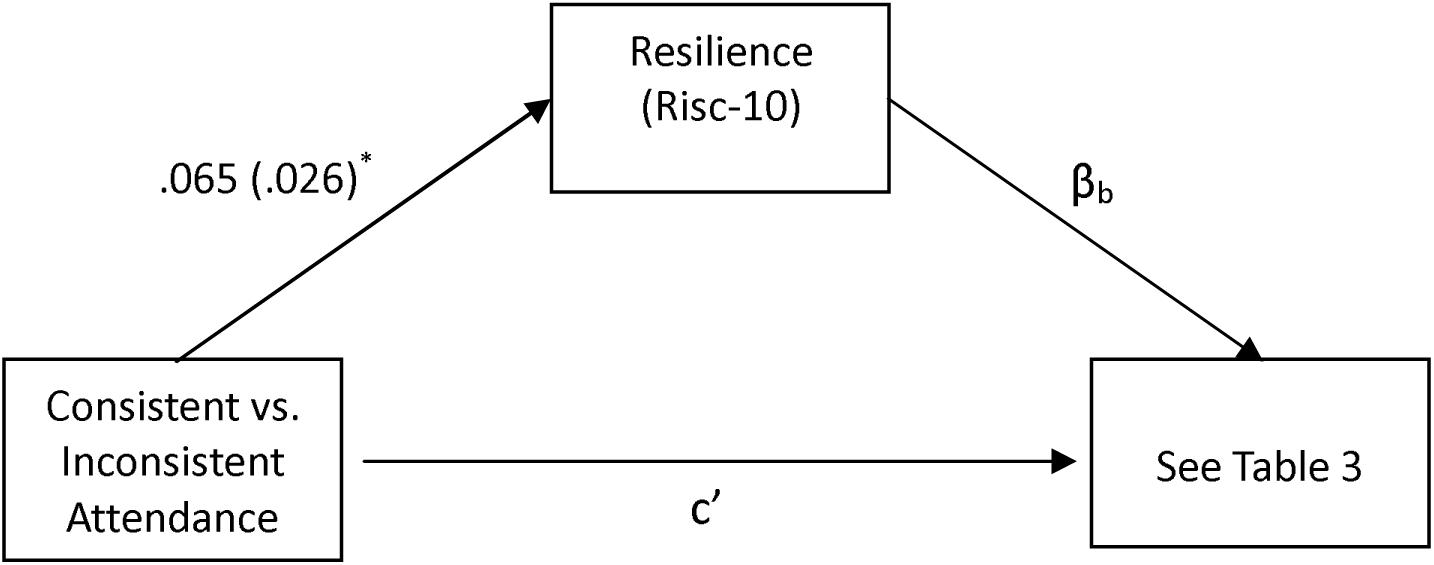
Mediational pathway between consistent vs. inconsistent attendance, resilience, and secondary outcomes

**Table 3:** Mediation Estimates.

| **Secondary Outcome** | **Total Effect** | **Direct Effect** | **Indirect Effect CI**<br>**(lower 95%,**<br>**upper 95%)** | **Mediation type** |
| --- | --- | --- | --- | --- |
| **Perceived stress** | (TE = -0.11, SE = .004, $P = 0.002$) | (DE = -0.06, SE = 0.03, $p = 0.03$) | (-0.12, -0.02)* | Partial |
| **Depression** | (TE = -0.06, SE = 0.04, $p = 0.09$) | (DE = -0.02, SE = 0.03, $p = 0.51$) | (-0.07, -0.01)* | Full |
| **Persist** | (TE = 0.06, SE = 0.03, $p = 0.04$) | (DE = 0.03, SE = 0.03, $p = 0.33$) | (0.007, 0.07)* | Full |
| **Self-efficacy** | (TE = 0.06, SE = 0.03, $p = 0.039$) | (DE = 0.04 SE = 0.02, $p = 0.11$) | (0.006, 0.08)* | Full |
| **Self-awareness: Proactive at Work** | (TE = 0.05, SE = 0.03, $p = 0.07$) | (DE = 0.01 SE = 0.22, $p = 0.69$) | (0.009, 0.07)* | Full |
| **Self-awareness: Reflective Self-Development** | (TE = 0.07, SE = 0.03, $p = 0.007$) | (DE = 0.04 SE = 0.02, $p = 0.12$) | (0.007, 0.07)* | Full |

**Table 4:** Path Estimates.

| Path | Estimate | SE | CI lower, upper 95% CI | p |
| --- | --- | --- | --- | --- |
| **(Consistent vs. inconsistent) attendance → Resilience** | 0.07 | 0.03 | (0.01, -0.12) | 0.01 |
| *Perceived stress* |  |  |  |  |
| **Resilience → Perceived stress** | -0.70 | 0.05 | (-0.79, -0.61) | <0.001 |
| **Attendance → Perceived stress** | -0.06 | 0.03 | (-0.12, -0.005) | 0.03 |
| *Depression* |  |  |  |  |
| **Resilience → Depression** | -0.58 | 0.05 | (-0.68, -0.48) | <0.001 |
| **Attendance → Depression** | -0.02 | 0.03 | (-0.09, -0.04) | 0.50 |
| *Persist* |  |  |  |  |
| **Resilience → Persist** | 0.57 | 0.04 | (0.49, 0.65) | <0.001 |
| **Attendance → Persist** | 0.03 | 0.03 | (-0.03, 0.08) | 0.33 |
| *Self-Efficacy* |  |  |  |  |
| **Resilience → Self-efficacy** | 0.65 | 0.03 | (0.58, 0.72) | <0.001 |
| **Attendance → Self-efficacy** | 0.04 | 0.02 | (-0.008, 0.08) | 0.11 |
| *Self-awareness: Proactive at work* |  |  |  |  |
| **Resilience → Proactive at work** | 0.63 | 0.03 | (0.57, 0.70) | <0.001 |
| **Attendance → Proactive at work** | 0.01 | 0.02 | (-0.04, 0.05) | 0.69 |
| *Self-awareness: Reflective Self-Development (RSD)* |  |  |  |  |
| **Resilience → RSD** | 0.57 | 0.03 | (0.50, 0.63) | <0.001 |
| **Attendance → RSD** | 0.04 | 0.02 | (-0.009, 0.08) | 0.12 |

### Aim 4: Program satisfaction and self-perceived changes on post-program survey

Most of the participants rated the program good to excellent overall (*M* = 4.43, *SD* = 0.75; 97.9% ≥ 3), found the program valuable (*M* = 4.60, *SD* = 0.68; 98.0% ≥ 3), and would recommend it to a friend or a colleague (*M* = 4.71, *SD* = 0.61; 98.5% ≥ 3). Almost all of these satisfaction scores hovered around the top of the scale point at 5. There was also a significant increase in self-reported knowledge of how to become a more resilient scientist compared to before the program, (*t*(610) = 51.39, *p* < 0.001; *d* = 2.08; *M*_change_ = 2.42, *SD*_change_ = 1.16).

As expected, compared to inconsistent attenders, consistent attenders reported significantly higher ratings of the BRS program, value, and were more likely to recommend the program to a colleague or a friend. Moreover, those who attended consistently were more likely to report that their perceived resilience had increased, that they gained important skills that help with school or home, and that they developed a greater ability to manage stress and conflict. However, we did not see a difference in their self-perceived ability to become a better scientist.

### Ancillary analyses: Impact of the trainee demographics

Because BRS had high proportion of trainees from diverse backgrounds, we explored whether there is a differential effect of BRS by race/ethnicity (dichotomized as white vs. non-white trainees, N=295 and 308, respectively). Although we did not find a significant differences in the pre-and-post matched measures on race/ethnicity, we found significant differences in self-perceived ratings in the post-survey for ethnicity. For example, two-way between group analyses of self-perceived resilience change (gaining important skills; the ability to manage stress and conflict; and the ability to become a better scientist) on race/ethnicity by attendance revealed no significant interactions (all ps >.1) but significant main effects (all ps<.01), albeit moderate to small effect sizes. The results are presented in Table 6 *(see S3 for additional breakdown of effects of BRS race/ethnicity).* There were no significant effects of gender.

**Table 5:**
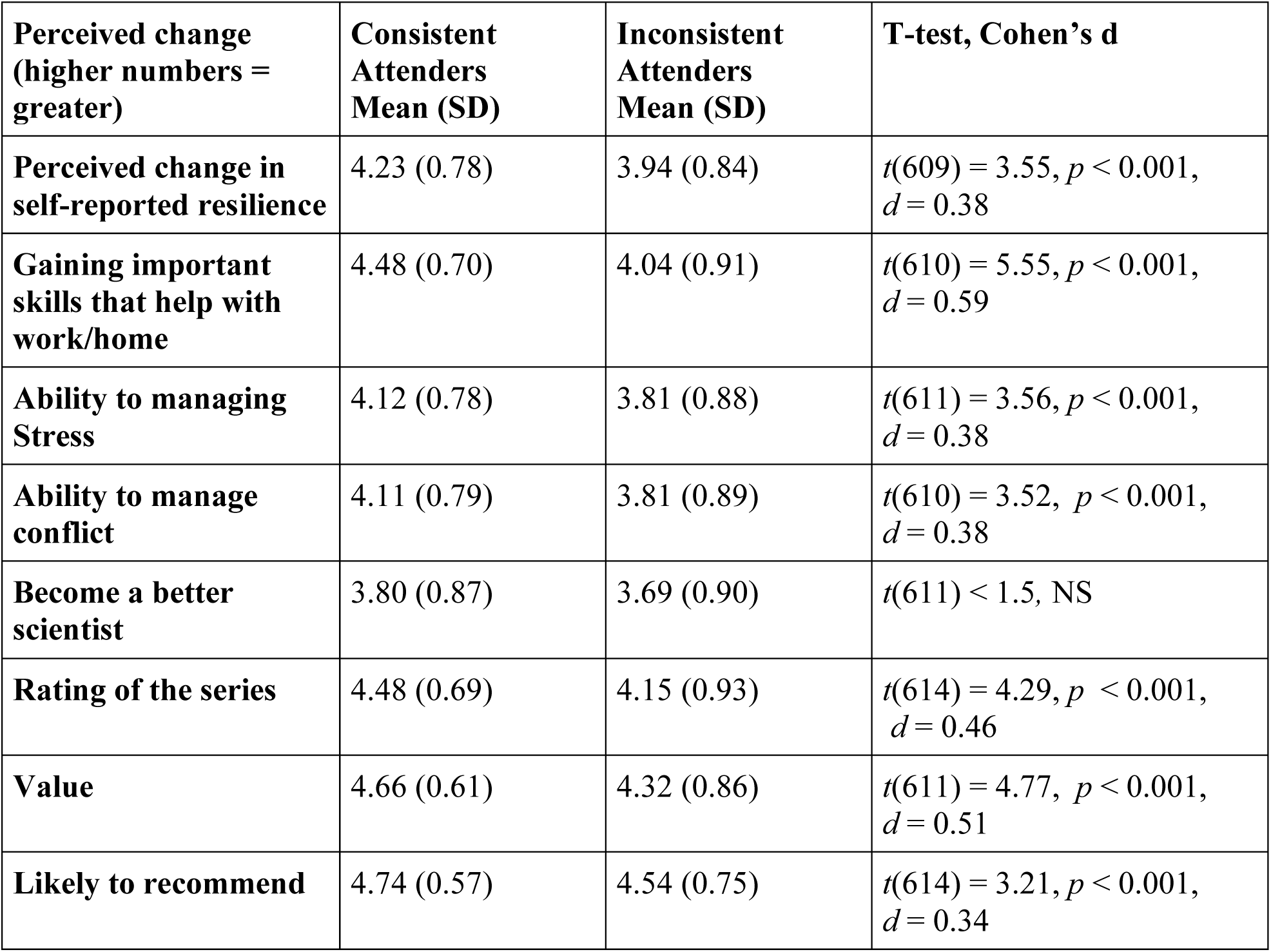
Self-perceived changes for consistent vs. inconsistent attenders.

| **Perceived change**<br>**(higher numbers =**<br>**greater)** | **Consistent**<br>**Attendees**<br>**Mean (SD)** | **Inconsistent**<br>**Attendees**<br>**Mean (SD)** | **T-test, Cohen's d** |
| --- | --- | --- | --- |
| **Perceived change in**<br>**self-reported resilience** | 4.23 (0.78) | 3.94 (0.84) | $t(609) = 3.55, p < 0.001, d = 0.38$ |
| **Gaining important**<br>**skills that help with**<br>**work/home** | 4.48 (0.70) | 4.04 (0.91) | $t(610) = 5.55, p < 0.001, d = 0.59$ |
| **Ability to managing**<br>**Stress** | 4.12 (0.78) | 3.81 (0.88) | $t(611) = 3.56, p < 0.001, d = 0.38$ |
| **Ability to manage**<br>**conflict** | 4.11 (0.79) | 3.81 (0.89) | $t(610) = 3.52, p < 0.001, d = 0.38$ |
| **Become a better**<br>**scientist** | 3.80 (0.87) | 3.69 (0.90) | $t(611) < 1.5, \text{NS}$ |
| **Rating of the series** | 4.48 (0.69) | 4.15 (0.93) | $t(614) = 4.29, p < 0.001, d = 0.46$ |
| **Value** | 4.66 (0.61) | 4.32 (0.86) | $t(611) = 4.77, p < 0.001, d = 0.51$ |
| **Likely to recommend** | 4.74 (0.57) | 4.54 (0.75) | $t(614) = 3.21, p < 0.001, d = 0.34$ |

**Table 6:** Self-perceived changes between White vs. Non-white trainees.

| Perceived change<br>(Higher numbers =<br>greater) | White trainees<br>Mean (SD) | Non-white<br>trainees<br>Mean (SD) | Main effect, Partial eta<br>squared ($\eta_p^2$) |
| --- | --- | --- | --- |
| Perceived change in<br>self-reported resilience | 4.04 (0.79) | 4.32 (0.75) | $F(1,596) = 19.03, p < 0.001,$<br>$\eta_p^2 = 0.031$ |
| Gaining important<br>skills that help | 4.35 (0.79) | 4.47 (0.71) | $F(1,597) = 9.60, p = 0.002,$<br>$\eta_p^2 = 0.016$ |
| Managing Stress | 4.00 (0.79) | 4.15 (0.79) | $F(1,598) = 5.68, p = 0.002,$<br>$\eta_p^2 = 0.010$ |
| Managing Conflict | 3.98 (0.82) | 4.16 (0.79) | $F(1,597) = 7.08, p = 0.008,$<br>$\eta_p^2 = 0.012$ |
| Become a Better<br>Scientist | 3.61 (0.85) | 3.96 (0.87) | $F(1,598) = 11.71, p < 0.001,$<br>$\eta_p^2 = 0.019$ |
|  |  |  | *(main effect of attendance is NS)* |

## Conclusions

The results show that the BRS is clearly effective as an intervention program with a moderate to large impact on the trainees. In addition to being rated highly, the program appears to increase resilience, self-efficacy, and self-awareness. For those who completed both pre-and-post program measures, BRS improved participants’ self-perceived ability to shift and persist, self-efficacy, and self-awareness while decreasing self-reported anxiety, depression, perceived stress, and presenteeism. After the intervention, participants reported that they were better managing stress and conflict, that they found what they learned valuable, and that they had become more resilient.

As expected, we saw greater effects among those who attend more sessions indicating that the program is effective and is especially beneficial for those who consistently attended (i.e., “higher dose over time”). In the post-program survey only sample, consistent attenders showed increased resilience, ability to persist, self-efficacy, proactive and reflective self-development components of self-awareness, and saw decreases in perceived stress and marginal decrease in depression. We found that the program’s impact of resilience fully mediated its effects on depression, ability to persist, self-efficacy, and self-awareness, and partially mediated perceived stress. This hints at the underlying mechanism that the increase in resilience is driving the corresponding changes on the secondary outcomes. We speculate that perceived stress may only be partially mediated by resilience, because BRS had another important component – a sense of community and that the trainees are not alone. In fact, in our comment section, the most frequently mentioned comments were that the trainees were glad that they are not alone or that their experiences are not unique. Part of the mechanism driving the reduction in perceived stress may not only be from the increase in resilience but also from a sense of relief or comfort that the experience is shared by others [see 39]. That sense of not being alone may also reduce feelings of shame which may further improve one’s ability to ask for help and work on building effective coping strategies. Furthermore, those who attended more than half also found the program more valuable and were more likely to recommend to their friend or colleague; however, it should be noted that those who attended less than half still rated the series highly, found it valuable, and likely to recommend it to a friend or a colleague.

Although BRS seems to be beneficial for all trainees taken together, it seems to have been especially beneficial for non-white trainees. Compared to white trainees, non-white trainees self-reported and perceived much greater changes in their perception of resiliency, gained important skills that help them in their work and home, learned to manage stress and conflict better, and reported that they become a better scientist as a result, despite not showing greater changes in resilience measures. We speculate that, similar to not being alone effect noted above, that nonwhite trainees may find comfort in knowing that their experiences and challenges are shared and recognized by others. BRS explicitly address the challenges that marginalized trainees face in science and research community – especially the role of bias, microaggression, stereotype threat, and how it can lead to attributional ambiguity [40]. These findings indicate that BRS could be an important and effective tool in retaining diverse trainees in biomedical science.

It appears that BRS can fill a critical void that currently exists in the biomedical training community. It is empowering trainees to effectively deal with the stressors of academics and research by giving them a sense of agency and the best strategies and tools to cope with those stressors, as well as setbacks and other adversities. It is a step towards reducing the mental health crisis amongst trainees in biomedical sciences, and when scaled up, could provide a large benefit, and help prevent attrition of trainees in science.

## Discussion

The BRS series is a program designed to help trainees struggling with stressors and raise their resilience during a time when the alarm bells started sounding regarding the graduate mental health crisis in science and when the pandemic was exacerbating that effect. The program was an attempt at meeting the crucial needs of the trainees, and helping them alleviate and address common stressors, providing them with coping skills, and ultimately increasing resilience for those pursuing biomedical science. And it appears that the program is successful at meeting those goals and is addressing the needs of the trainees and improving their well-being.

However, some may wonder if the beneficial changes in this intervention program were mainly driven by the changes in the pandemic stressors and time. The BRS1 started before the COVID-19 vaccine was available and at the height of the period during the pandemic when trainees were grappling with uncertainty and isolation and ended during the potentially more hopeful period of the pandemic when vaccines started becoming available. Although this alternative explanation cannot be ruled out completely, if this were the case, BRS2 should not show the same results. BRS2 started as the vaccines were becoming widely available and the pressures of the pandemic had started easing. Yet we did not see any differences between BRS1 and BRS2. This strongly suggests that the BRS intervention effectiveness is not just due to the effects of the pandemic (i.e., effects of history) and that it is more likely that the intervention is targeting the needs of the trainees.

Another alternative explanation for the result could be that the participants who were consistent attenders were inherently different at baseline. In order to rule out this explanation, we conducted pre-program measure differences between consistent vs. inconsistent attenders on all primary and secondary measures. We did not find any significant pre-measure differences (all ps>.2). Therefore, it appears that both consistent and inconsistent attenders were equivalent on our primary and secondary measures at the baseline (at the beginning of the program) and the program attendance or dosage effect is seemingly driving the effect. On the other hand, we cannot say for sure that those who attended all of the sessions and those who missed one or more were identical in all possible ways. Given the severity of the pandemic, including widespread isolation, salient racial injustice issues, and concerns for various other mental health issues of the trainees at that time, a decision was made to make the program open for all trainees who wanted to participate. Hence, a critical decision was made to forgo a control comparison condition (i.e., waitlist control) and, therefore, drop the quasi-experimental design to allow for broad and open participation. A more ethical decision was made at the expense of establishing the causal impact of the intervention program, and we are unable to rule out some confounding variables which underscores the need for cautious interpretation. Thus, acknowledging this weakness, future studies might compare the efficacy of this intervention with that of other interventions or introduce a waitlist control.

While we utilized a well-known and validated survey instruments for this study, we had to make compromises in the interest of survey length and administration time. We made a deliberate choice to utilize only a subset of questions on some instruments and calculate means for the sake of survey length and administration time. This decision may have implications for the psychometric properties of the measures. While the subset of questions retained their essential reliability and validity, it is possible that the overall robustness of the measures may have been affected, as they were originally designed to be considered as a whole. This limitation underscores the need for caution when interpreting the results, particularly in instances where a more comprehensive evaluation of psychometric properties are evaluated. However, these limitations were necessary to ensure practicality and participation in our study, and we are confident that it does not distract from the overall effectiveness of the program.

Although the program is largely successful, there was significant attrition. In both rounds of BRS, we started with approximately 600 trainees, and by the end of session, we had approximately 250 trainees in attendance. We speculate that there are two reasons for this. First is the time commitment and pressure from others to work while in the lab during the workday. As the attrition occurred, small group facilitators informally reached out to a subset of trainees, querying why they stopped attending. Many trainees mentioned that the time commitment interfered with lab work commitments, and they chose to prioritize lab work. Some said their PI/supervisor did not support them attending a webinar during the day when they should be focused on lab work. It is essential to acknowledge that the impact of this time commitment and the pressure to prioritize lab work may introduce a selection bias in our study. It is possible that our sample may not fully represent trainees who are heavily or overcommitted to lab work, those who prioritize it over self-care/improvement, or those who lack support for their well-being and resilience from their PIs/supervisors. However, we contend that this underscores the potential benefits of mandating resilience training as a vital component of the curriculum. This step is crucial as we strive to foster a cultural shift in the field of science. Many trainees were hesitant to commit three to six hours per month to improving their resilience because of fears that participation would interfere with their lab work. We need to change the culture of long hours and complete dedication to lab work to also emphasizing the importance of self-care and self-improvement. After all, one must be well to do well.

The second possibility for attrition is motivation. Although many trainees were excited to start, they may have lacked the motivation and commitment to complete the entire series. Hence, it is possible that only the motivated trainees completed the program and benefited from it. Nevertheless, this explanation for attrition does not undermine the effectiveness of the BRS program. The social psychological literature on behavior and attitude change state that one needs to be willing (i.e., motivated) and able first before any actual changes can occur (e.g., theory of planned behavior) [41]. The effectiveness of BRS is likely driven by providing the tools and the skills the trainees need to implement and make the changes. Those who are willing and able are more likely to practice those skills and see corresponding changes in their resilience. Our data hint that this could be the case – those who attend more than half of the sessions (likely more motivated and committed) report more benefits than those who attend less than half. Furthermore, our pre-and-post sample group demonstrated the greatest positives changes in all secondary outcomes, and they were also the group who attended the majority the BRS sessions. In fact, while we saw changes in anxiety, work presenteeism, and the ability to shift during stressful times in our pre-and-post matched-sample, we did not find the corresponding changes between consistent and inconsistent attenders on post-program only sample. It is possible that these variables require consistent effort and motivation to improve. Hence, it is unlikely that any intervention program, no matter how effective, will have a large impact on those who are unmotivated and unwilling to change.

The BRS has demonstrated its effectiveness as an intervention, and it could prove to be a valuable tool for trainees as they navigate the unique challenges of academic scientific settings. Furthermore, the program is readily available to a wide range of trainees since it is free of charge and easy to participate in. Trainees have the option of joining live sessions twice a year with small discussion groups via Zoom, or they can watch recordings at any time at their convenience. The program’s affordability and accessibility are particularly advantageous to trainees who are constrained by financial and scheduling limitations. Given the program’s accessibility, there is little reason why trainees and extramural institutes should not explore the possibility of incorporating it in their training (see *S3*, on how OITE can aid in BRS adaptation at various extramural institutions).

The BRS program has been shown to be effective in enhancing resilience among trainees, and especially beneficial for individuals from diverse backgrounds. By providing trainees with the tools to manage stress, cope with failure, and maintain a healthy work-life balance, the program has the potential to retain a talented biomedical workforce while nurturing a group of resilient future scientists. Considering the mental health crisis that many biomedical and science trainees face, the BRS program may be an important component of addressing these issues in the sciences more broadly. With its proven effectiveness, accessibility, and potential to improve trainee well-being, the BRS program offers a promising solution to some of the challenges facing the scientific community.

## Data Availability

All data produced in the present study are available upon reasonable request to the authors

## Acknowledgments

This paper is dedicated to the memory of Wes Beckstead (1983-2011) who greatly influenced the work of SLM in developing support structures for NIH intramural trainees.

We thank Drs. Laura Koehly, Janetta Lun, Brett Pelham, and the OITE staff for valuable input on this project, and Dr. Michael Gottesman for supporting the development of well-being programs for the NIH intramural scientists. We appreciate the on-going support of the NIH Immediate Office of the Director, NIH Office of Research on Women’s Health, NIH Sexual and Gender Minority Research Office, NIH Office of AIDS Research, NIH Office of Behavioral and Social Sciences Research, and other NIH Institutes and Centers. Finally, we are deeply grateful for the energy and enthusiasm of the facilitators, mentors, and trainees who participated in BRS and who continue to influence the work of the NIH OITE through their participation and feedback.

## Supporting Information

S1 file. BRS sessional titles and descriptions

S2 file. Self-perceived changes by specific race/ethnicity

S3 file. How OITE can aid in BRS adaptation at various extramural institutions

## References

1. Frank A. Science: It’s Really, Really Hard, And That’s Something To Celebrate. National Public Radio. 2012 Feb. Available from: https://www.npr.org/sections/13.7/2012/02/14/146857164/science-its-really-really-hard-and-thats-something-to-celebrate

2. Drew C. Why science majors change their mind: it’s just so darn hard. The New York Times. 2011 Nov 4. Available from: https://www.nytimes.com/2011/11/06/education/edlife/why-science-majors-change-their-mind-its-just-so-darn-hard.html?_r=2

3. Coe R. Comparability of GCSE examinations in different subjects: an application of the Rasch model. Oxford Review of Education. 2008;34(5):609–636. doi:10.1080/03054980801970312

4. Abbott A. Huge survey reveals pressures of scientist’s lives. Nature, 2020; 577(7791), 460–461.

5. Evans TM, Bira L, Gastelum JB, Weiss LT, Vanderford NL. Evidence for a mental health crisis in graduate education. Nat Biotechnol. 2018;36(3):282. doi:10.1038/nbt.4089

6. Woolston C. PhDs: the tortuous truth. Nature. 2019 Nov;575(7782):403-406. doi: 10.1038/d41586-019-03459-7.

7. Matulevicius SA, Kho KA, Reisch J, Yin H. Academic medicine faculty perceptions of work-life balance before and since the COVID-19 pandemic. JAMA Netw Open. 2021;4(6):e2113539. doi:10.1001/jamanetworkopen.2021.13539

8. Chan C, Oey NE, Tan EK. Mental health of scientists in the time of COVID-19. Brain Behav Immun. 2020; 88:956. doi:10.1016/j.bbi.2020.05.039

9. Mattijssen L, Van Vliet N, Van Doorn T, Kanbier N, Teelken C. PNN PhD Survey: Asking the relevant questions. Mental wellbeing, workload, burnout, research environment, progress of the PhD project, considering to quit. Promovendi Netwerk Nederland; 2020.

10. Grogan KE. How the entire scientific community can confront gender bias in the workplace. Nat Ecol Evol. 2019;3(1):3–6. doi:10.1038/s41559-018-0747-4

11. Maher MA, Wofford AM, Roksa J, Feldon DF. Exploring early exits: Doctoral attrition in the biomedical sciences. J Coll Stud Retent. 2020;22(2):205–226. doi:10.1177/1521025117736871

12. Flahtery C. Mental Health Crisis for Grad Students. Inside Higher Ed. 2018. Available from: https://www.insidehighered.com/news/2018/03/06/new-study-says-graduate-students-mental-health-crisis

13. Joyce S, Shand F, Tighe J, Laurent SJ, Bryant RA, Harvey SB. Road to resilience: a systematic review and meta-analysis of resilience training programmes and interventions. BMJ Open. 2018;8(6):e017858. doi:10.1136/bmjopen-2017-017858

14. Thoman SE, DiBona T, Abelar J, Robnett RD. STEMing from scholarship and resilience: A case study focusing on US undergraduate women who are thriving in STEM. Int J Gend Sci Technol. 2020;12(1):122–151.

15. Maher MA, Wofford AM, Roksa J, Feldon DF. Exploring early exits: Doctoral attrition in the biomedical sciences. Journal of College Student Retention: Research, Theory & Practice. 2020 Aug;22(2):205–26.

16. Diekman AB, Steinberg M, Brown ER, Belanger AL, Clark EK. A Goal Congruity Model of Role Entry, Engagement, and Exit: Understanding Communal Goal Processes in STEM Gender Gaps. Personality and Social Psychology Review. 2017;21(2):142–175. doi:10.1177/1088868316642141

17. American Psychological Association. Building your resilience. APA. 2020 Feb 1. Available from: https://www.apa.org/topics/resilience/building-your-resilience

18. Cohn MA, Fredrickson BL, Brown SL, Mikels JA, Conway AM. Happiness unpacked: Positive emotions increase life satisfaction by building resilience. Emotion. 2009;9(3):361–368. doi:10.1037/a0015952

19. Fredrickson B, Cohn M, Coffey K, Pek J, Finkel S, Judd CM. Open hearts build lives: Positive emotions, induced through loving-kindness meditation, build consequential personal resources. J Pers Soc Psychol. 2008;95(5):1045–1062. doi:10.1037/a0013262

20. Mak WWS, Ng ISW, Wong CCY. Resilience: Enhancing well-being through the positive cognitive triad. J Couns Psychol. 2011;58:610–617. doi:10.1037/a0025195

21. Thogersen-Ntoumani C, Black J, Lindwall M, Whittaker A, Balanos G. Presenteeism, stress resilience, and physical activity in older manual workers: A person-centred analysis. Eur J Ageing. 2017;14(4):385–396. doi:10.1007/s10433-017-0418-3

22. Seligman M, Schulman P, Sarason IG. Explanatory style as a predictor of productivity and quitting among life insurance sales agents. J Pers Soc Psychol. 1986;50(4):832–838. doi:10.1037/0022-3514.50.4.832

23. Van Katwyk PT, Fox S, Spector PE, Kelloway EK. Using the Job-Related Affective Well-being Scale (JAWS) to investigate affective responses to work stressors. J Occup Health Psychol. 2000;5:219–230. 10.1037/1076-8998.5.2.219

24. Kim-Cohen J. Resilience and developmental psychopathology. Child Adolesc Psychiatr Clin N Am. 2007;16(2):271–283. doi:10.1016/j.chc.2006.11.003

25. Forbes S, Fikretoglu D. Building resilience: The conceptual basis and research evidence for resilience training programs. Rev Gen Psychol. 2018;22(4):452–468. doi:10.1037/gpr0000152

26. Robertson I, Cooper C, Sarkar M, Curran T. Resilience training in the workplace from 2003 to 2014: A systematic review. J Occup Organ Psychol. 2015;88(3):533–562. doi:10.1111/joop.12120

27. Youssef CM, Luthans F. Positive organizational behavior in the workplace: The impact of hope, optimism, and resilience. J Manage. 2007;33(5):774–800. 10.1177/0149206307305562

28. Johnson JR, Emmons HC, Rivard RL, Griffin KH, Dusek JA. Resilience training: a pilot study of a mindfulness-based program with depressed healthcare professionals. Explore (NY). 2015;11(6):433–444. doi:10.1016/j.explore.2015.08.002

29. Connor KM, Davidson JR. Development of a new resilience scale: The Connor-Davidson resilience scale (CD-RISC). Depression and anxiety. 2003;18(2):76–82. doi:10.1002/da.10113

30. Cohen S, Kamarck T, Mermelstein R. A global measure of perceived stress. J Health Soc Behav. 1983:385–396. doi:10.2307/2136404

31. Spitzer RL, Kroenke K, Williams JB, Löwe B. A brief measure for assessing generalized anxiety disorder: the GAD-7. Arch Intern Med. 2006;166(10):1092–1097. doi:10.1001/archinte.166.10.1092

32. Kroenke K, Spitzer RL, Williams JB. The PHQ-9: validity of a brief depression severity measure. J Gen Intern Med. 2001;16(9):606–613. doi:10.1046/j.1525-1497.2001.016009606.x

33. Gilbreath B, Frew EJ. The stress-related presenteeism scale [measurement instrument]. Pueblo (CO): Hasan School of Business, Colorado State University; 2008.

34. Chen E, McLean KC, Miller GE. Shift-and-persist strategies: Associations with socioeconomic status and the regulation of inflammation among adolescents and their parents. Psychosom Med. 2015;77(4):371–382. doi:10.1097/PSY.0000000000000157

35. Chen G, Gully SM, Eden D. Validation of a new general self-efficacy scale. Organizational Research Methods. 2001;4(1):62–83. doi:10.1177/109442810141004

36. Sutton A. Measuring the effects of self-awareness: Construction of the Self-Awareness Outcomes Questionnaire. Eur. J. Psychol. 2016;12(4):645. doi:10.5964/ejop.v12i4.1178

37. Preacher KJ, Hayes AF. SPSS and SAS procedures for estimating indirect effects in simple mediation models. Behav Res Methods Instrum Comput. 2004;36:717–731. doi:10.3758/BF03206553

38. Preacher KJ, Rucker DD, Hayes AF. Addressing moderated mediation hypotheses: Theory, methods, and prescriptions. Multivariate behavioral research. 2007 Jun 29;42(1):185–227.

39. Neff KD. Self-compassion: An alternative conceptualization of a healthy attitude toward oneself. Self Identity. 2003;2:85–101.

40. Crocker J, Voelkl K, Testa M, Major B. Social stigma: The affective consequences of attributional ambiguity. J Pers Soc Psychol. 1991;60(2):218–228. doi:10.1037/0022-3514.60.2.218

41. Ajzen I. The theory of planned behavior. Organizational behavior and human decision processes. 1991;50(2):179–211. doi:10.1016/0749-5978(91)90020-T

